# Magnetic seizure therapy and electroconvulsive therapy increase aperiodic activity

**DOI:** 10.1101/2023.01.11.23284450

**Authors:** Sydney E. Smith, Eena L. Kosik, Quirine van Engen, Jordan Kohn, Aron T. Hill, Reza Zomorrodi, Daniel M. Blumberger, Zafiris J. Daskalakis, Itay Hadas, Bradley Voytek

**Affiliations:** Neurosciences Graduate Program; Department of Cognitive Science; Herbert Wertheim School of Public Health and Human Longevity Science; Department of Psychiatry, School of Medicine, University of California, San Diego, La Jolla, CA, USA; Cognitive Neuroscience Unit, School of Psychology, Deakin University, Geelong, Victoria, Australia; Department of Psychiatry, Central Clinical School, Monash University, Melbourne, Australia; Temerty Centre for Therapeutic Brain Intervention, Centre for Addiction and Mental Health, University of Toronto, Toronto, Ontario, Canada; Halıcıoğlu Data Science Institute; Kavli Institute for Brain and Mind, University of California, San Diego, La Jolla, CA, USA

**Author notes:** ^#^These authors contributed equally (Sydney E. Smith, Eena L. Kosik, Quirine van Engen, Itay Hadas, Bradley Voytek).

## Abstract

Major depressive disorder (MDD) is a leading cause of disability worldwide. One of the most efficacious treatments for treatment-resistant MDD is electroconvulsive therapy (ECT). Recently, magnetic seizure therapy (MST) was developed as an alternative to ECT due to its more favorable side effect profile. While these approaches have been very successful clinically, the neural mechanisms underlying their therapeutic effects are unknown. For example, clinical “slowing” of the electroencephalogram beginning in the postictal state and extending days to weeks post-treatment has been observed in both treatment modalities. However, a recent longitudinal study of a small cohort of ECT patients revealed that, rather than delta oscillations, clinical slowing was better explained by increases in aperiodic activity, an emerging EEG signal linked to neural inhibition. Here we investigate the role of aperiodic activity in a cohort of patients who received ECT and a cohort of patients who received MST treatment. We find that aperiodic neural activity increases significantly in patients receiving either ECT or MST. Although not directly related to clinical efficacy in this dataset, increased aperiodic activity is linked to greater amounts of neural inhibition, which is suggestive of a potential shared neural mechanism of action across ECT and MST.

## Introduction

Since its development in 1938, electroconvulsive therapy (ECT) has been used as a treatment for mood disorders including Major Depressive Disorder (MDD) and particularly, treatment-resistant depression (TRD)^1^. During a session of ECT, an electrical current is applied to the scalp of an anesthetized patient which induces a seizure as it passes through the brain. Despite the remission rates between 50-70%^2^, it remains one of the least used treatments for depression. Fewer than 1% of patients with TRD receive ECT due to a combination of fear, stigma, and concerns about cognitive side effects, such as short-term amnesia^3^. The search for alternative, yet comparably effective, therapeutic stimulation techniques has led to the development of treatments like repetitive transcranial magnetic stimulation (rTMS) and more recently, magnetic seizure therapy (MST).

MST is a more focal treatment that was developed to mimic the therapeutic effects of ECT while minimizing the adverse side effects. Specifically, MST involves the application of a magnetic field to produce a seizure in the brain. The first person to receive MST was treated in 2000^4^. Compared to ECT, MST can produce remission rates between 30-60%^5–7^ and patients receiving MST experience fewer cognitive side effects and recover more quickly after the procedure compared to patients receiving ECT^5,8–10^(**Fig. 1**). ECT and MST differ in the characteristics of the induced seizure, with the ECT-induced seizure spreading to deeper subcortical structures, including the hippocampus, while the MST-induced seizure is more confined to the cortex^11–14^. These differences in ictal expression, along with the differences in the electric vs. magnetic fields, are assumed to underlie the distinct side effect profiles of the two treatments^9,14,15^. Despite the clinical efficacy of both forms of treatment, their neural mechanism(s) of action remain unclear. Candidate mechanisms include the anticonvulsant hypothesis, wherein seizure threshold increases with subsequent treatments^16^, and the neurotrophic hypothesis, which posits that ECT induces neuroplastic processes in various brain structures, particularly the hippocampus^17^. However, these hypotheses are difficult to test in humans. Here, we leverage a putative biomarker for neural excitation/inhibition that is derived from non-invasive electroencephalography (EEG) to test the hypothesis that the therapeutic effects of ECT and MST may arise from a similar neural mechanism of action.

**Fig. 1.**
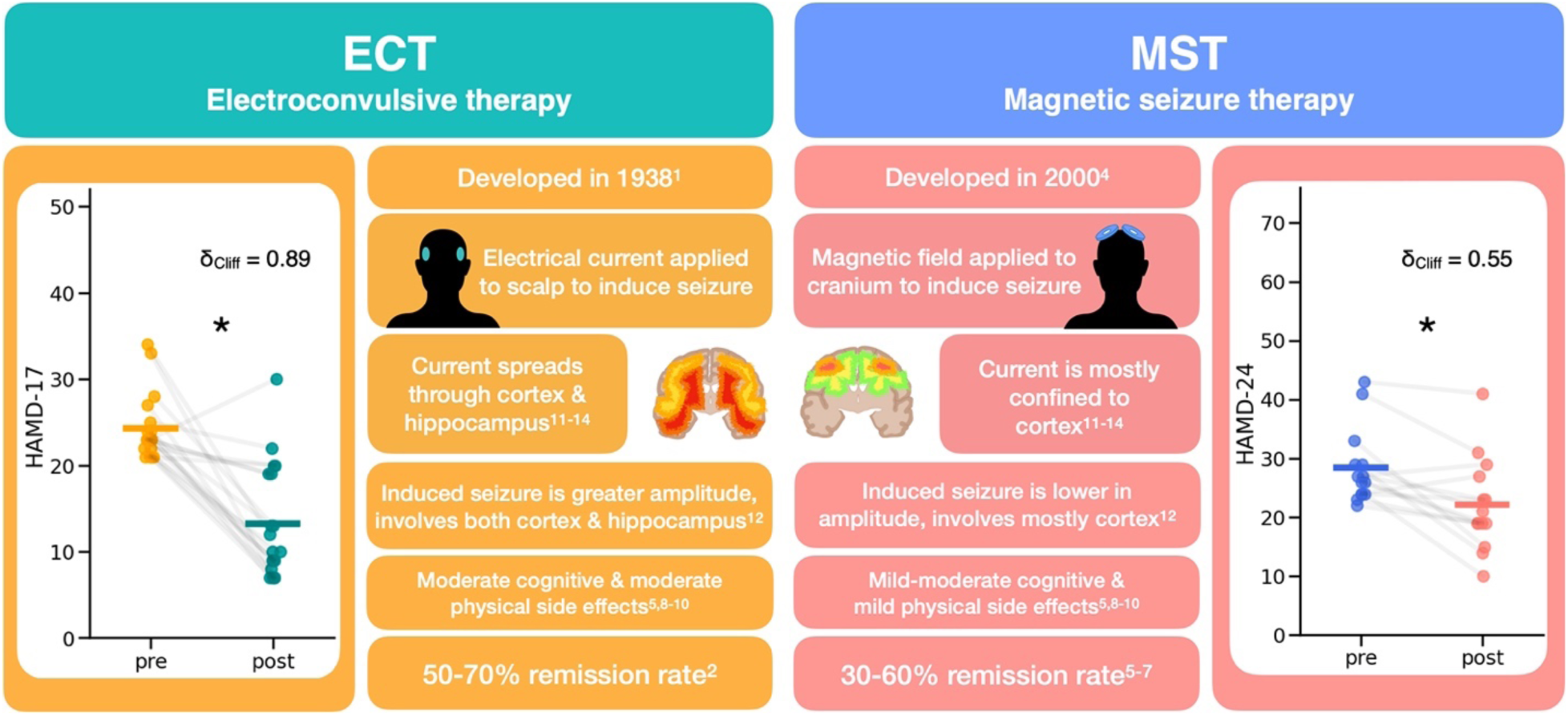
Overview of ECT and MST. This describes important details of both treatment types, electroconvulsive therapy (ECT), and magnetic seizure therapy (MST). Both treatments are typically only used on patients with treatment-resistant depression and involve inducing a seizure, either with an electrical current or a magnetic field. The main difference is that ECT has a more global spread to subcortical structures and hippocampus, whereas MST affects more local cortical structures. However, both treatment types significantly reduce depression ratings, with MST having a comparable but more modest therapeutic effect than ECT. We can see this clinical improvement in the datasets analyzed here,, as measured by the HAMD-17 for ECT (pre median(IQR) = 23.0 (22, 24.5), post median(IQR) = 10 (8.5, 19), W(18) = 6, δ_Cliff_ = 0.89, p = 5.3 x 10^-5^) and the HAMD-24 for MST (pre-MST = 26.5 (24, 29), post-MST = 20 (19, 26), W(13) = 7, δ_Cliff_ = 0.55, p = 2.3 x 10^-3^).

Although its relationship to therapeutic efficacy is disputed, post-treatment EEG recordings both in patients receiving ECT and those receiving MST are characterized by clinical “slowing.” This effect is quantified as increased spectral power in the delta (1-4 Hz) and theta (4-8 Hz) frequency ranges compared to baseline^18,19^. These changes persist for days to weeks following treatment cessation and are consistent between MST and ECT^18–20^. Sometimes, this increase in spectral power can be observed as apparent high amplitude delta and theta oscillations in the time domain^21,22^. However, this slowing has not been consistently linked to any clear therapeutic mechanism of action or clinical efficacy for either treatment modality^23^, illustrating an important gap in understanding of EEG signatures underlying clinical response to convulsive therapy.

One possibility for the persistent ambiguity is that previous investigations of band power changes have not considered the contributions of aperiodic activity to EEG signals. This is important because traditional analysis methods conflate periodic (oscillatory) activity with aperiodic activity^24,25^. For example, spectral power at any frequency is often assumed to represent sustained oscillatory activity, when in actuality the signal more frequently contains only short bursts of neural oscillations^26–28^. Even when no oscillations are present, a large aperiodic signal can appear very similar to slowing in the EEG when measuring spectral power, as we have recently demonstrated^29^. This effect occurs because the EEG signal is a mix of oscillations and aperiodic activity, where oscillations are defined by concentrated power within a specific, narrow frequency band. In contrast, aperiodic activity manifests as a broadband phenomenon, where power decreases exponentially as a function of frequency (1/f*^χ^*scaling). This exponential relationship is parameterized by *χ* (**Fig. 2A**), which naturally arises from the physiology of the EEG signal^30,31^. The large increase in low frequency power following an increase in the aperiodic exponent, or a “steepening” of the power spectrum, is often mistaken as an increase in the actual power of slow oscillations (**Fig. 2B-C**). **Figure 2D** provides a visual representation of the difference in between these two situations in the time domain.

**Fig. 2.**
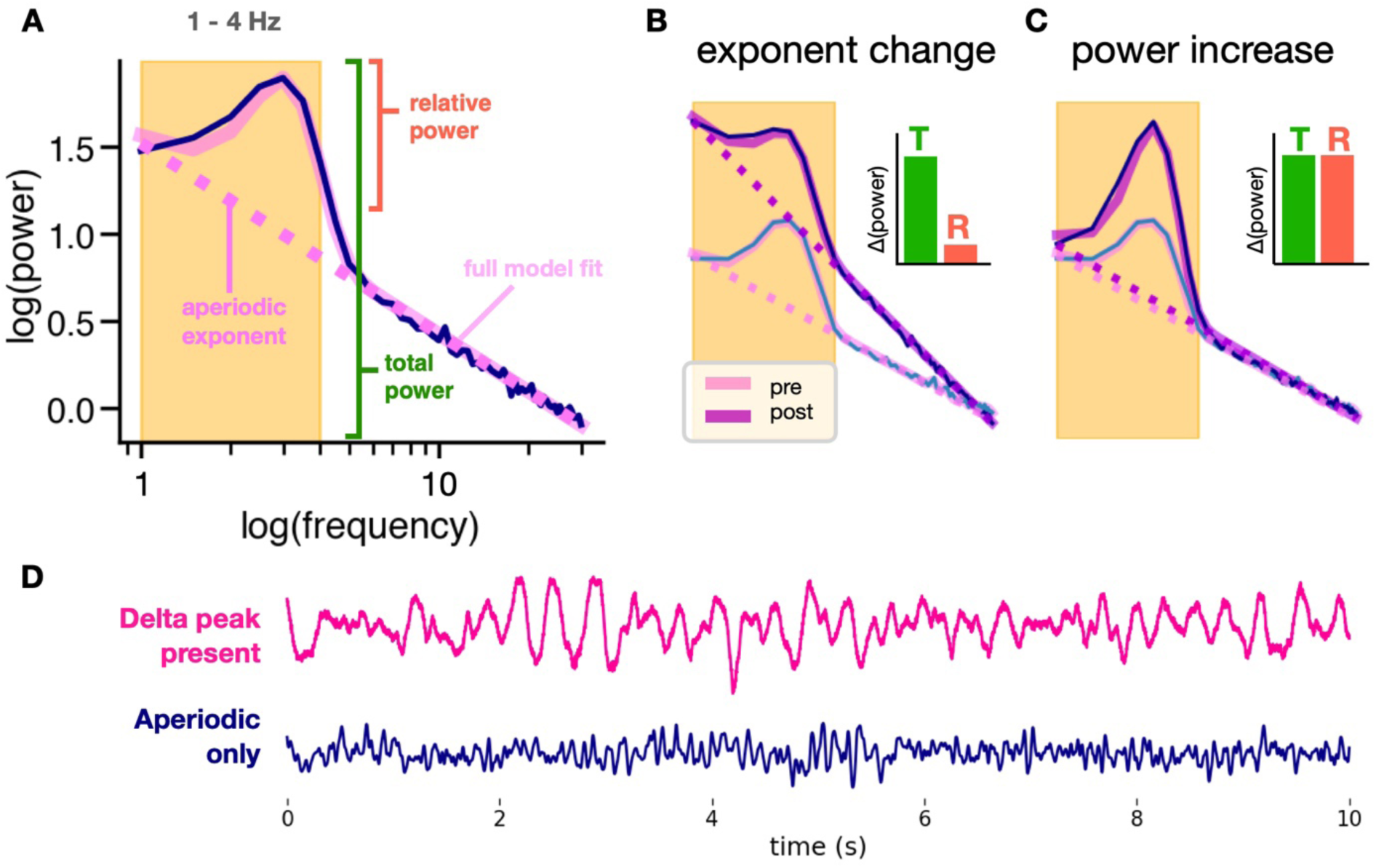
Using spectral parameterization to disambiguate periodic and aperiodic contributions to delta band power. **(A)** Simulated power spectrum illustrating parameterized spectra. Unlike traditional band power measures that conflate periodic and aperiodic activity, spectral parameterization defines oscillation power as relative power above the aperiodic component (pink dashed line). **(B)** Increases in the aperiodic exponent can cause apparent increases in total (T) band power, while power relative (R) to the aperiodic component remains unchanged. We see this here in a simulated power spectrum depicting an increase in exponent with no delta oscillation changes after treatment. **(C)** True increases in oscillation power show increases in both total power and relative power. We see this here in a simulated power spectrum depicting an increase in delta oscillation power after treatment with no change in exponent. **(D)** Delta in the EEG trace vs. aperiodic activity. EEG with delta oscillations (where a delta peak is present in the spectra) is visibly different from EEG with only aperiodic activity in the delta band.

An increase in aperiodic activity and thereby, inhibition, could be a possible physiological mechanism underlying the efficacy of these treatments. For instance, the aperiodic exponent has been shown to at least partially capture the relative excitatory and inhibitory contributions to the local field potential^32,33^. When the spectrum steepens, reflected in an increase in the aperiodic exponent, this corresponds to a shift toward greater inhibitory tone. In the power spectrum, this manifests as a large increase in low frequency power with a concomitant decrease in high frequency power. In the context of ECT and MST, an increase in the aperiodic exponent, thereby a relative increase in inhibition, is in line with the cortical inhibition theory of depression, which posits that patients with MDD have insufficient inhibitory activity in various brain regions^34^.

Our recent investigation into longitudinal changes in aperiodic activity throughout a course of ECT revealed that many of the observations of increased frontal delta band power could be better explained by increases in frontal aperiodic activity^29^. Specifically, in that report, we found that the aperiodic exponent significantly increased longitudinally throughout the course of ECT treatment. Furthermore, both aperiodic exponent at baseline and the magnitude of the change in exponent throughout treatment were related to treatment response, as measured by the Quick Inventory of Depressive Symptomatology - Self Report (QIDS-SR). In comparison, there were no longitudinal effects in delta band power or actual delta oscillation power after accounting for the broadband aperiodic signal. This was interpreted as evidence that longitudinal increases in frontal aperiodic activity may better explain clinical slowing observed in seizure-inducing treatments than slow oscillations.

Here, we sought to replicate and extend that smaller (n = 9), longitudinal study in a larger sample of two previously collected independent datasets, one from a study of patients receiving ECT (n = 22)^35^ and the other from a registered clinical trial of patients receiving MST (n = 23)^36^(Clinicaltrials.gov, NCT01596608). The analyses described here are retrospective, since these datasets have already finished data collection, however, potential changes in aperiodic activity had not yet been investigated in either study. We hypothesized that our recent discovery of increased frontal aperiodic activity in a longitudinal study of ECT would generalize to interventional findings of slowing in both ECT and MST in these acqurired datasets^18,19^. To test this hypothesis we compare measures of aperiodic activity, oscillations, and canonical band power. For each treatment modality, resting-state EEG was collected at baseline and after completing a standard treatment. We find that in both ECT and MST treatment conditions, aperiodic exponent increases significantly, representing a putative increase in inhibitory tone. This study presents promising opportunities for future research to better understand and differentiate the mechanisms of clinical efficacy for both ECT and MST.

## Results

### Clinical effects

For patients that received ECT treatment and had complete pre- and post-ECT clinical ratings (n=19), the severity of depressive symptomatology as assessed by the clinician using the Hamilton Depression Rating Scale (HAMD-17) decreased significantly (pre-ECT median(IQR) = 23 (22, 24.5), post-ECT median(IQR) = 10 (8.5, 19), W(18) = 6, δ_Cliff_ = 0.89, p = 5.3 x 10^-5^) (**Fig. 1**, left, and **Table 1**). For patients that received MST treatment and had complete pre- and post-MST clinical ratings (n = 15), HAMD-24 scores also decreased significantly (pre-MST = 26.5 (24, 29), post-MST = 20 (19, 26), W(13) = 7, δ_Cliff_ = 0.55, p = 2.3 x 10^-3^) (**Fig. 1**, right, and **Table 1**). Because different versions of the HAM-D were collected when the ECT study and MST clinical trial were conducted, comparison of clinical efficacy between the two treatments should be interpreted with caution.

**Table 1.** ECT and MST dataset details for patients included in analysis. ECT dataset details were from Voineskos et al., 2016^35^ and further correspondence. MST dataset details were from Daskalakis et al., 2020^36^ and further correspondence. Remission rate was calculated as a percentage of the included patients in our analysis from the cohort that showed equal or larger than 50% decrease in symptom severity.

|  | **ECT:** | **MST:** |
| --- | --- | --- |
| **N** | 22 | 22 |
| **Age (median (IQR))** | 46.5 (32, 61) | 47.5 (40.75, 52) |
| **Gender (Male/Female)** | 8/12 (2 unknown) | 11/11 |
| **HAM-D pre (median (IQR))** | 23 (22, 24) (HAMD-17) | 26.5 (24, 29) (HAMD-24) |
| **HAM-D post (median (IQR))** | 10 (8.5, 19) (HAMD-17) | 20 (19, 26) (HAMD-24) |
| **Number of treatments received (median (IQR))** | 14 (10, 17.5) | 24 (18, 24) |
| **Remission rate** | 63% | 21% |
| **Medications** |  |  |
| **Antidepressant** | 89% | 65% |
| **Antipsychotic** | 42% | 25% |
| **Anxiolytics** | 32% | 15% |
| **Stimulants** | 11% | 15% |
| **Sedative** | 68% | 35% |

### Treatment-related EEG effects: ECT

Because eight different tests of EEG features were run on the same sample of patients receiving ECT, a Holm-Bonferroni correction was applied (see **Supp. Table 1**). Compared to baseline, patients who received a full course of ECT exhibited significant increases in aperiodic activity, as measured by the aperiodic exponent of the EEG power spectra, which become visibly steeper (pre = 0.88 ± 0.21 µV^2^Hz^-1^, post = 1.25 ± 0.33 µV^2^Hz^-1^, t(21) = -9.07, d_z_ = 2.00, ɑ_adj_ = 6.25 x 10^-3^, p = 1.05 x 10^-8^) (**Fig. 3A-B**). We also observed concomitant increase in delta band power (pre = -11.92 ± 0.31 µV^2^Hz^-1^, post = -11.03 ± 0.58 µV^2^Hz^-1^, t(21) = -8.30, d_z_ = 1.88, ɑ_adj_ = 7.14 x 10^-3^, p = 4.83 x 10^-8^) (**Fig. 3C**). Furthermore, we observed a small but significant change in delta oscillation power, which was adjusted for aperiodic activity (pre = 0.16 (0.08, 0.66) µV^2^, post = 0.46 (0.23, 0.76) µV^2^, W(11) = 10, δ_Cliff_ = -0.26, ɑ_adj_ = 5.00 x 10^-2^, p = 0.02). Importantly, only 12 out of 22 patients had detectable delta oscillation peaks both at baseline and after ECT, which casts doubt on the idea that observed clinical slowing is solely representative of an increase in existing slow wave oscillatory activity. In this case, slowing could also be driven by a change in aperiodic activity post-ECT (**Fig. 3D**). Alternatively, the emergence of delta oscillations might also contribute to observations of clinical slowing. To measure this, we computed delta abundance, defined as the fraction of electrodes exhibiting a delta oscillation peak in their power spectra, above the aperiodic signal. We found that delta abundance increases significantly post-ECT (pre = 0.023 (0, 0.15), post = 0.36 (0.07, 0.67), W(21) = 20.5, δ_Cliff_ = -0.67, ɑ_adj_ = 1.00 x 10^-2^, p = 1.77 x 10^-4^) (**Fig. 3E**).

**Fig. 3.**
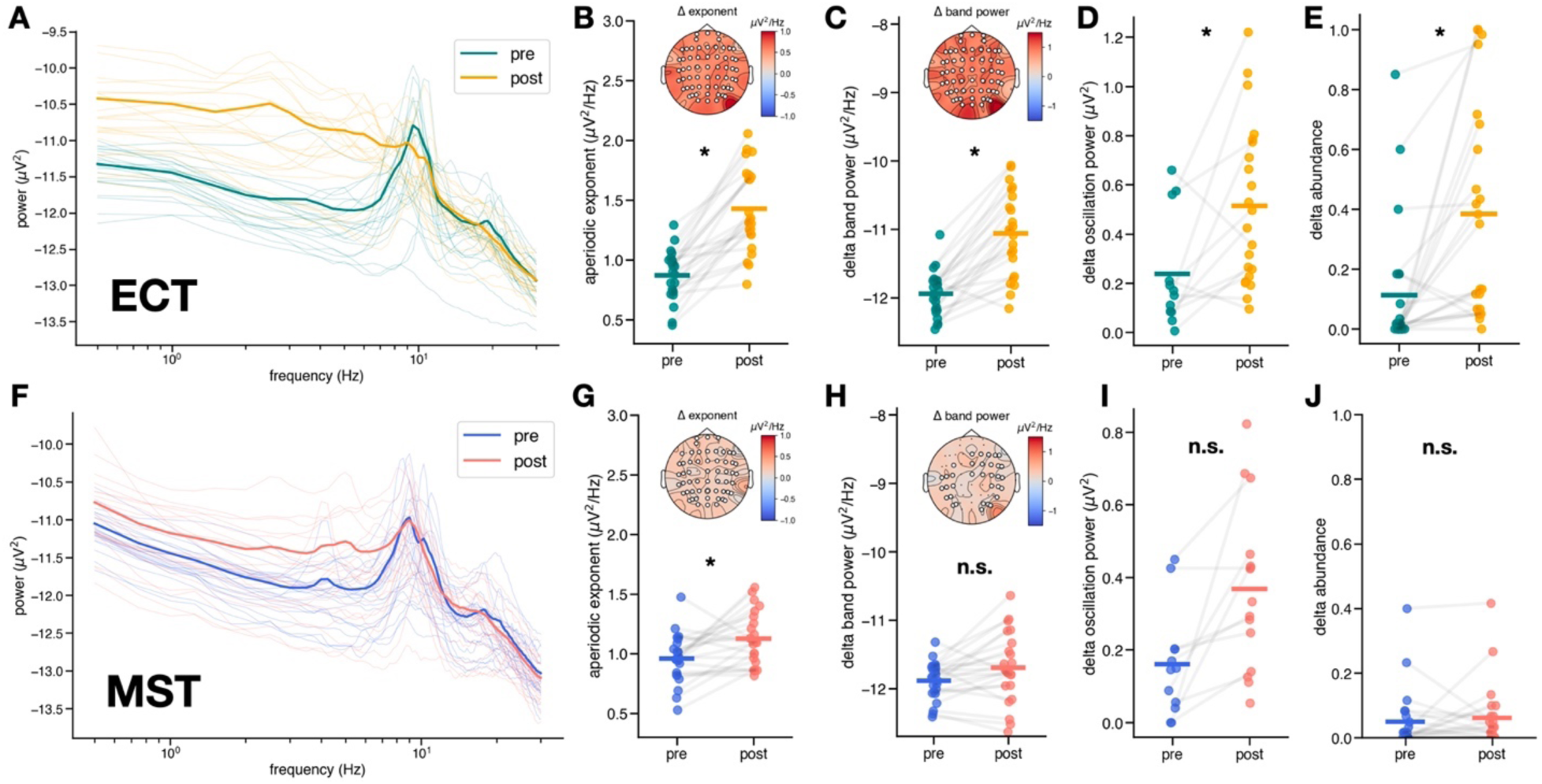
EEG results - Aperiodic vs. delta band power slowing. Spectral differences in aperiodic exponent and delta oscillations in ECT (top) and MST (bottom). **(A)** Raw power spectra averaged across channels for each patient pre- and post-ECT. Bolded spectra represent average across patients. **(B)** Increase in aperiodic exponent post-ECT (pre = 0.88 ± 0.21 µV^2^Hz^-1^, post = 1.25 ± 0.33 µV^2^Hz^-1^, t(21) = -9.07, d_z_ = 2.00, ɑ_adj_ = 6.25 x 10^-3^, p = 1.05 x 10^-8^), inset shows scalp topography of median exponent change, with significant electrodes (p < 0.05) marked in white. **(C)** Increase in total power in the delta band post-ECT (pre = -11.88 ± 0.27 µV^2^Hz^-1^, post = -11.69 ± 0.51 µV^2^Hz^-1^, t(21) = -2.23, d_z_ = 0.45, ɑ_adj_ = 1.25 x 10^-2^, p = 0.036), inset shows scalp topography of median delta band power change, with significant electrodes (p < 0.05) marked in white. **(D)** Increase in aperiodic-adjusted oscillation power in the delta band – only 12 out of 22 patients exhibited a delta oscillation peak both pre- and post-ECT (pre = 0.16 (0.08, 0.66) µV^2^, post = 0.46 (0.23, 0.76) µV^2^, W(11) = 10, δ_Cliff_ = -0.26, ɑ_adj_ = 5.00 x 10^-2^, p = 0.02). Many patients exhibited an emergence of delta peaks post-ECT, hence the increased number of data points in post-ECT. No scalp topography is depicted because delta oscillation presence was not consistent across electrodes and patients. **(E)** Increase in the abundance of delta oscillations post-ECT (pre = 0.023 (0, 0.15), post = 0.36 (0.07, 0.67), W(21) = 20.5, δ_Cliff_ = -0.67, ɑ_adj_ = 1.00 x 10^-2^, p = 1.77 x 10^-4^). **(F)** Raw power spectra averaged across channels for each patient pre- and post-MST. Bolded spectra represent average across patients. **(G)** Increase in aperiodic exponent post-MST (pre = 0.98 ± 0.18 µV^2^Hz^-1^, post = 1.14 ± 0.21 µV^2^Hz^-1^, t(21) = -3.06, d_z_ = 0.80, ɑ_adj_ = 7.14 x 10^-3^, p = 6.0 x 10^-3^), inset shows scalp topography of median exponent change, with significant electrodes (p > 0.05) marked in white. **(H)** No significant change in total power in the delta band post-MST (pre = -11.88 ± 0.27 µV^2^Hz^-1^, post = -11.69 ± 0.51 µV^2^Hz^-1^, t(21) = -2.23, d_z_ = 0.45, ɑ_adj_ = 1.25 x 10^-2^, p = 0.036), inset shows scalp topography of median delta band power change, with significant electrodes (p > 0.05) marked in white. **(I)** No significant change in aperiodic-adjusted oscillation power in the delta band–only 10 out of 22 patients exhibited a delta oscillation peak both pre- and post-MST (pre = 0.16 ± 0.14 µV^2^, post = 0.35 ± 0.21 µV^2^, t(9) = -3.26, d_z_ = 1.14, ɑ_adj_ = 8.33 x 10^-3^, p = 9.8 x 10^-3^), with a few patients exhibiting emerging delta peaks post-MST, hence the increased number of data points post-MST. **(J)** No significant change in the abundance of delta oscillations post-MST (pre = 0.02 (0, 0.07), post = 0.03 (0, 0.05), W(21) = 62.5, δ_Cliff_ = -0.15, ɑ_adj_ = 5.00 x 10^- 2^, p = 0.80).

To further investigate how much of the observed changes in delta band power, or slowing, were driven by aperiodic activity compared to delta oscillations, we performed a multiple linear regression to examine the contributions of delta oscillation power, delta abundance, and aperiodic activity to delta band power. Note that the ɑ-thresholds for main effects were adjusted according to Holm-Bonferroni multiple comparison correction. For the 12 patients whose power spectra contained oscillation peaks both pre- and post-ECT, this regression was significant overall (R^2^_adj_ = 0.63, F(3, 8) = 7.32, p = 0.01), with the aperiodic exponent contributing more substantially to observations of increased delta band power (β = 0.69, ɑ_adj_ = 1.67 x 10^-2^, p = 0.02, 95% CI[0.11, 1.26]) compared to delta abundance (β = 0.12, ɑ_adj_ = 0.05, p = 0.64, 95% CI[-0.46, 0.70]), or delta oscillation power (β= 0.39, ɑ_adj_ = 2.50 x 10^-2^, p = 0.07, 95% CI[-0.04, 0.83]), although this trending result is not significant under a Holm-Bonferroni correction. That is, traditional band power definitions of delta, which do not seek to examine whether or not true oscillations are present, might be better explained by non-oscillatory aperiodic activity in this sample. Furthermore, we performed similar multiple linear regressions to see whether theta and alpha band power are more significantly related to the change in aperiodic exponent, or the change in abundance of the respective frequency bands. Full details of these results are in the supplementary materials.

We also observed changes in oscillation power in the theta and alpha ranges in patients who received ECT (**Fig. 4C**). For the 20 patients whose EEG power spectra contained theta oscillation peaks before and after treatment, we observed significant increases in aperiodic-adjusted theta oscillation power (pre = 0.30 ± 0.15 µV^2^, post = 0.70 ± 0.32 µV^2^, t(19) = -5.65, d_z_ = 1.55, ɑ_adj_ = 8.33 x 10^-3^, p = 1.90 x 10^-5^) (**Fig. 4A**). Similar to what we observed for delta, the fraction of electrodes that exhibited a theta oscillation peak (theta abundance) also increased significantly with ECT (pre = 0.23 (0.03, 0.63), post = 0.69 (0.34, 0.91), W(21) = 35, δ_Cliff_ = -0.45, ɑ_adj_ = 1.67 x 10^-2^, p = 5.40 x 10^-3^) (**Fig. 4B**). When we repeated this analysis in the alpha band, we found that alpha oscillation power decreased post-ECT compared to baseline (pre = 1.32 ± 0.49 µV^2^, post = 0.99 ± 0.39 µV^2^, t(21) = 3.33, d_z_ = 0.78, ɑ_adj_ = 1.25 x 10^-2^, p = 3.20 x 10^-3^) as did alpha abundance (pre = 1.0 (1, 1), post = 1.0 (0.94, 1), W(21) = 42 δ_Cliff_ = 0.35, ɑ_adj_ = 2.50 x 10^-2^, p = 0.020 (**Fig. 4** **D-E**). This result is notable because alpha band power increases post-ECT when measured using the canonical bandpass approach (pre = -11.59 ± 0.58 µV^2^Hz^-1^, post = -11.38 ± 0.36 µV^2^Hz^-1^, t(21) = -2.06, d_z_ = 0.41, p = 0.05), highlighting the importance of spectral parameterization to disambiguate the contributions of periodic and aperiodic activity to band power, which separates band power into the periodic and aperiodic components.

**Fig. 4.**
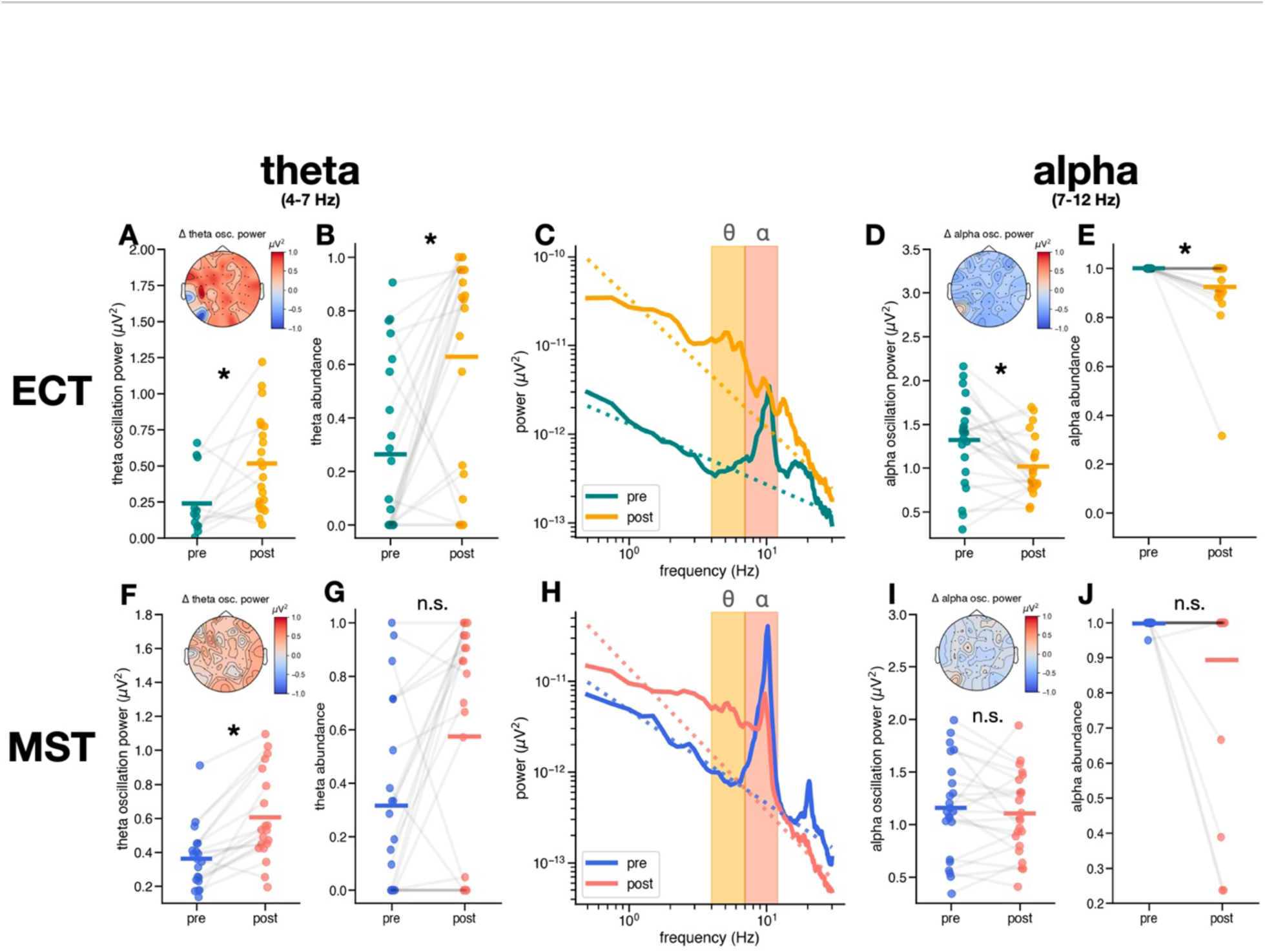
EEG results – Changes in theta and alpha oscillations. Changes in theta (4-7 Hz) and alpha (7-12 Hz) oscillations in ECT (top) and MST (bottom). **(A)** Observed increase in theta oscillation power post-ECT (pre = 0.30 ± 0.15 µV^2^, post = 0.70 ± 0.32 µV^2^, t(19) = -5.65, d_z_ = 1.55, ɑ_adj_ = 8.33 x 10^-3^, p = 1.90 x 10^-5^), inset shows scalp topography of median theta oscillation change. **(B)** Increase in theta abundance theta abundance post-ECT (pre = 0.23 (0.03, 0.63), post = 0.69 (0.34, 0.91), W(21) = 35, δCliff = -0.45, ɑ_adj_ = 1.67 x 10^-2^, p = 5.40 x 10^-3^). **(C)** Power spectra from electrode F8 in a patient who received ECT showing the emergence of a theta oscillation and a decrease in alpha oscillation power post-ECT. **(D)** Increase in alpha oscillation power post-ECT (pre = 1.32 ± 0.49 µV^2^, post = 0.99 ± 0.39 µV^2^, t(21) = 3.33, d_z_ = 0.78, ɑ_adj_ = 1.25 x 10^-2^, p = 3.20 x 10^-3^), inset shows scalp topography of median alpha oscillation power change. **(E)** Decrease in alpha abundance post-ECT (pre = 1.0 (1, 1), post = 1.0 (0.94, 1), W(21) = 42 δCliff = 0.35, ɑadj = 2.50 x 10^-2^, p = 0.020). **(F)** Increase in theta oscillation power post-MST (pre = 0.35 (0.14, 0.42) µV^2^, post = 0.53 (0.44, 0.82) µV^2^, W(17) = 3.0, δ_Cliff_ = -0.97, ɑ_adj_ = 6.25 x 10^-3^, p = 3.80 x 10^-5^), inset shows scalp topography of median theta oscillation power change. **(G)** No significant change in theta abundance (pre = 0.39(0.07, 0.98), post = 0.68(0.21, 0.95), W(21) = 47, δ_Cliff_ = - 0.34, ɑ_adj_ = 1.00 x 10^-2^, p = 0.02). **(H)** Power spectra from electrode F8 in a patient who received MST showing the emergence of a theta oscillation and a decrease in alpha oscillation power post-MST. **(I)** There is no significant change in alpha oscillation power post-MST (pre = 1.19 ± 0.44 µV^2^, post = 1.13 ± 0.37 µV^2^, t(21) = 0.88, d_z_ = 0.15, ɑ_adj_ = 2.50 x 10^-2^, p = 0.39), inset shows scallop topography of median change in alpha oscillation power. **(J)** No significant change in alpha abundance post-MST (pre = 1.0 (1.0, 1.0), post = 1.0 (1.0, 1.0), W(21) = 2.0, δ_Cliff_ = 0.18, ɑ_adj_ = 1.67 x 10^-2^, p = 0.18).

### Treatment-related EEG effects: MST

Similar to ECT, eight different tests of EEG features were run on the same sample of patients receiving MST, a Holm-Bonferonni correction was applied (see **Supp. Table 1**). We found that patients treated with MST (see Methods) also exhibited a significant increase in aperiodic exponent compared to baseline (pre = 0.98 ± 0.18 µV^2^Hz^-1^, post = 1.14 ± 0.21 µV^2^Hz^-1^, t(21) = -3.06, d_z_ = 0.80, ɑ_adj_ = 7.14 x 10^-3^, p = 6.0 x 10^-3^) (**Fig. 3G**). Although an increase in delta band power is trending, we did not observe significant changes with multiple comparison correction (pre = -11.88 ± 0.27 µV^2^Hz^-1^, post = -11.69 ± 0.51 µV^2^Hz^-1^, t(21) = - 2.23, d_z_ = 0.45, ɑ_adj_ = 1.25 x 10^-2^, p = 0.036), which would have been hypothesized under previous observations of clinical slowing in MST^16^. There was a trending increase in delta oscillation power for the 10 patients out of 22 whose spectra exhibited delta oscillation peaks both before and after MST (pre = 0.16 ± 0.14 µV^2^, post = 0.35 ± 0.21 µV^2^, t(9) = -3.26, d_z_ = 1.14, ɑ_adj_ = 8.33 x 10^-3^, p = 9.8 x 10^-3^), although this result was not significant under Holm-Bonferroni correction. Nor did we find a significant change in delta abundance post-MST compared to baseline in all patients (pre = 0.02 (0, 0.07), post = 0.03 (0, 0.05), W(21) = 62.5, δ_Cliff_ = -0.15, ɑ_adj_ = 5.00 x 10^-2^, p = 0.80) (**Fig. 3** **H, J**). However, the multiple linear regression to relate changes in delta band power to changes in aperiodic exponent, delta oscillation power, and delta abundance was significant overall (R^2^_adj_ = 0.68, F(3, 6) = 7.47, p = 0.02). In this model, increases in aperiodic activity (β = 0.89, ɑ_adj_ = 1.67 x 10^-2^, p = 6.22 x 10^-3^, 95% CI[0.36, 1.42]) better explain increases in delta band power than do changes in delta abundance (β = 0.09, ɑ_adj_ = 2.50 x 10^-2^, p = 0.71, 95% CI[-0.50, 0.69]) or oscillation power (β= -0.18, ɑ_adj_ = 0.05, p = 0.44, 95% CI[-0.73, 0.37]), indicating that aperiodic activity is likely driving observed clinical slowing in MST as well. Note that the ɑ-thresholds have been adjusted for multiple comparisons according to Holm-Bonferroni correction. Similar to our ECT analysis, we performed multiple linear regressions to relate theta and alpha band power to the change in aperiodic exponent, the change in abundance of the respective frequency bands, and their oscillation power. Full details of these results are in the supplementary materials.

Although we did not observe significant changes in delta abundance or delta oscillations post-MST, there were significant changes in theta oscillations (**Fig. 4H**). For the 20 out of 22 patients who exhibited theta oscillations, theta oscillation power increased significantly post-MST (pre = 0.35 (0.14, 0.42) µV^2^, post = 0.53 (0.44, 0.82) µV^2^, W(17) = 3.0, δ_Cliff_ = -0.97, ɑ_adj_ = 6.25 x 10^-3^, p = 3.80 x 10^-5^). However, for all patients, there was no significant change in theta abundance post-MST (pre = 0.39(0.07, 0.98), post = 0.68(0.21, 0.95), W(21) = 47, δ_Cliff_ = -0.34, ɑ_adj_ = 1.00 x 10^-2^, p = 0.02) (**Fig. 4** **F-G**). Furthermore, we observed no significant changes in alpha oscillation power (pre = 1.19 ± 0.44 µV^2^, post = 1.13 ± 0.37 µV^2^, t(21) = 0.88, d_z_ = 0.15, ɑ_adj_ = 2.50 x 10^-2^, p = 0.39), nor in alpha abundance post-MST (pre = 1.0 (1.0, 1.0), post = 1.0 (1.0, 1.0), W(21) = 2.0, δ_Cliff_ = 0.18, ɑ_adj_ = 1.67 x 10^-2^, p = 0.18) (**Fig. 4** **I-J**).

### Clinical improvement and the spectral features in ECT and MST

To investigate whether any of the spectral features at baseline or their changes post-ECT and -MST treatment could be related to a shared mechanism of action, we deployed an exhaustive research algorithm in combination with multiple linear regression. Specifically, we performed an exhaustive search to best relate post-treatment outcome, quantified using a normalized HAM-D score across the ECT and MST datasets. In this search, we accounted for methodological differences between datasets and treatment types by adding a fixed effect of which treatment the patient received. Normalized pre-treatment HAM-D score was also included as a fixed effect to include baseline clinical severity in the model. Considering EEG features for aperiodic activity, delta, and theta oscillations, exhaustive search identified baseline aperiodic exponent as most robustly related to treatment outcome (See Methods). Upon implementing this model in a multiple linear regression, the overall model was not significant (R^2^_adj_ = 0.04, F(3, 29) = 1.28, p = 0.30). Neither treatment type (β = 0.40, p = 0.44, 95% CI[-0.65, 1.45]), nor baseline clinical severity (β = 0.25, p = 0.34, 95% CI[-0.28, 0.76]) were significantly related to treatment outcome, whereas baseline exponent was trending (β = 0.30, p = 0.091, 95% CI[-0.05, 0.65]) (**Fig. 5**). This model should be interpreted with extreme caution, since the overall model was not significant, and the effect was trending. However, in summary, this relationship suggests that patients who begin treatment with smaller aperiodic exponents—or “flatter” spectra—tend to exhibit a less severe clinical profile after treatment, as measured by post-treatment HAM-D.

**Fig. 5.**
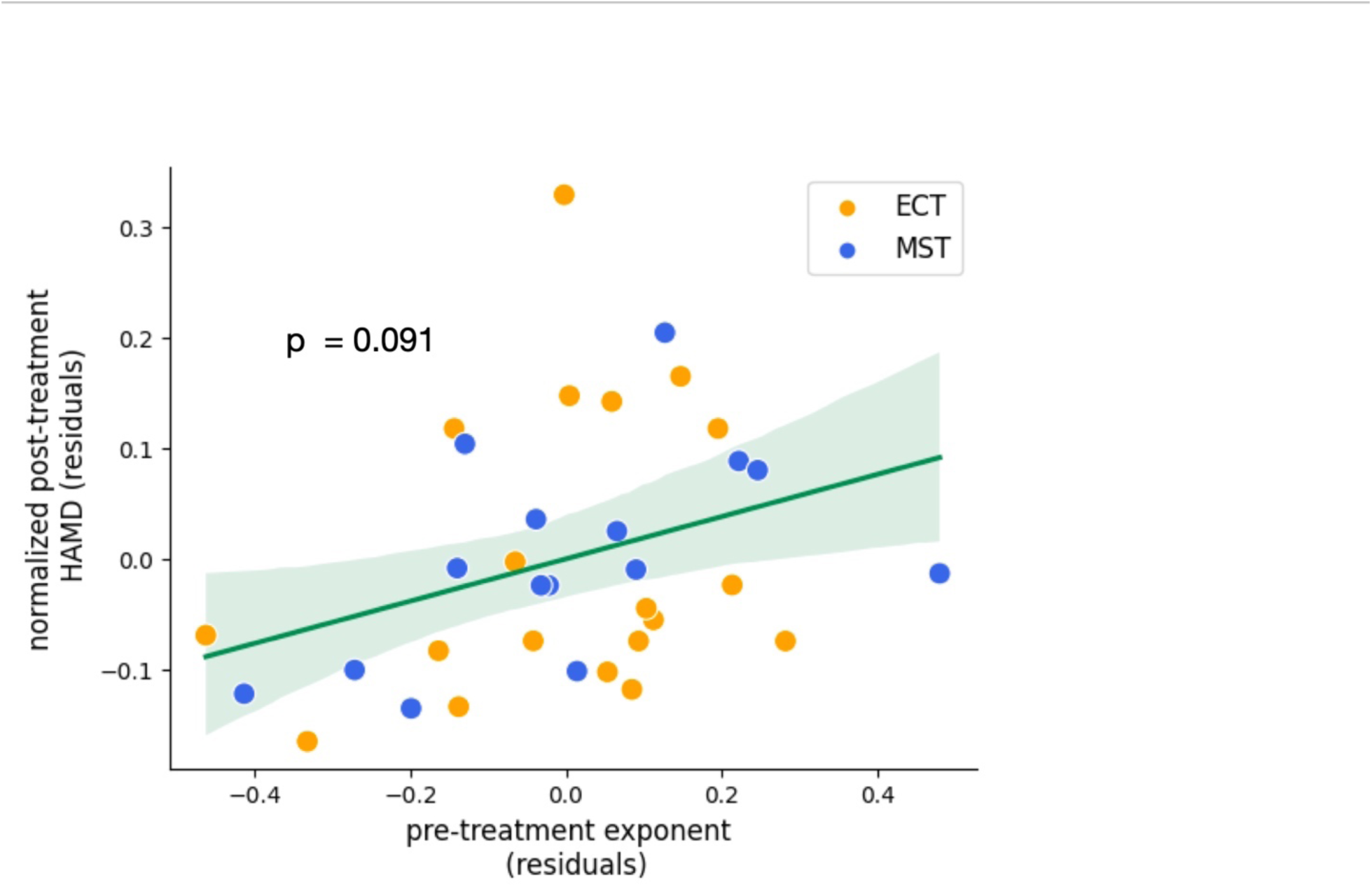
Partial regression analysis – baseline exponent and treatment outcome. Partial regression of combined ECT and MST datasets showing a positive trending relationship between patients’ aperiodic exponent at baseline and clinical outcome, as measured by normalized HAMD-D (β = 0.30, p = 0.091, 95% CI[-0.05, 0.657]). Here, patients whose baseline aperiodic exponent is lower, visible in a flatter pre-treatment power spectrum, show lower post-treatment symptom severity.

## Discussion

In this study, we investigated the hypothesis that changes in aperiodic activity might be related to a shared therapeutic mechanism of action for ECT and MST treatment. We observed that aperiodic activity increases after either ECT or MST. Although this finding supports the theory that aperiodic activity could be a potentially informative physiological change shared by both of these seizure-based treatments for depression, we found no direct support for a relationship between the magnitude of observed changes in aperiodic activity and clinical outcome. However, at the level of spectral changes, this increase in aperiodic activity is a more parsimonious explanation for observations of clinical slowing than delta band power or delta oscillations for both ECT and MST. These results replicate our recent finding from a smaller, longitudinal study in only ECT patients^29^.

Aperiodic activity has been widely associated with behavioral and disease states, such as cognitive and perceptual task performance^37–40^, development^41^, aging^42^, anesthesia^43^, ADHD^44^, and schizophrenia^45^. Furthermore, changes in aperiodic activity, like those observed in these two populations with MDD, have been associated with the physiological effects of deep brain stimulation as a treatment for MDD^46^. It is hypothesized that these aperiodic changes in the brain are related to the balance of excitation (E) and inhibition (I) based on simple computational models of the local field potential^32^, complex microcircuit models^47^, and experimental manipulations of EI balance using optogenetics^33^. The changes in aperiodic activity seen in patients undergoing ECT and MST, specifically increases in aperiodic exponent visible as a “steepening” of the power spectrum, are associated with relative increases in inhibitory activity. Increasing levels of inhibition as measured via the aperiodic exponent, align with observations of the anticonvulsant effects of ECT, with seizure induction threshold progressively increasing throughout a course of treatment1^6^.

Therapeutic interventions that potentially increase inhibitory activity are particularly relevant in light of the cortical inhibition theory of depression. According to this theory, patients with MDD have insufficient inhibitory activity^34^. Post-mortem tissue analyses have revealed that these patients have pathologically reduced numbers of inhibitory, GABAergic neurons^48^. Specifically, these patients have reduced somatostatin-expressing (SST) interneurons in prefrontal and limbic cortices^49–51^. Simulated biophysical models of human microcircuits with reduced SST activity produce LFP signals with flatter power spectra and lower aperiodic exponents compared to control simulations of microcircuits with healthy SST populations^52^. Deficits in these inhibitory interneuron populations could cause pathological dysfunction in EI balance in many areas, including prefrontal cortices. Because prefrontal cortices play an essential role in regulating EI balance throughout distributed networks in the brain^53^, dysfunctional inhibition in prefrontal regions could lead to widespread disruptions, including in limbic structures^54^ and the serotonergic and noradrenergic systems targeted by antidepressant medications^34^. Moreover, transcranial magnetic stimulation (TMS) concurrent with EEG also demonstrates decreased activation localized to frontal regions after successful neuromodulatory treatments for depression, and this attenuation is attributed to strengthening of inhibitory circuits^15,55^. Moreover, other physiological measure that are linked to EI balance, such as regional cerebral bloodflow (rCBF) or cerebral metabolic rater (rCMR)^55–58^, have been shown to change in response to ECT. Future work should compare these measures to the aperiodic signal to further investigate the physiological basis on an increase in exponent. In summary, increases in aperiodic exponent in patients receiving ECT and MST might reflect an increase in relative amounts of inhibitory activity, restoring pathologically low levels of inhibition to a healthier range.

Despite being linked to a promising mechanism of action, the lack of evidence for a direct relationship between increases in aperiodic exponent and the therapeutic effects of ECT and MST in this study presents a strong limitation. Although aperiodic exponent increases simultaneously as depression symptoms improve, the magnitude of these changes are unrelated in this sample, so aperiodic changes cannot be directly interpreted as a therapeutic mechanism of action for either treatment at this point, at least as measured by the HAM-D. This study was also limited by the lack of thorough evaluation for cognitive and physical side effects, which we believe present a fascinating and important opportunity for future investigation. Previous studies indicate that the magnitude of therapeutic effects of ECT are unrelated to the severity of cognitive side effects^59^, suggesting that these two phenomena are potentially dissociable. For instance, some theorists propose that the therapeutic mechanisms of ECT are related to increases in delta power—an effect we argue is better explained by increases in aperiodic activity— and that cognitive side effects are driven mostly by theta power^60^. The differences we observed in the patterns of significant results between ECT and MST in theta, as well as in delta and alpha oscillations, provide promising avenues for future investigation into the differential cognitive effects of these treatments. Theta oscillations, classically linked to memory^61^, are of special interest here. The difference between ECT and MST in the statistical significance of post-treatment emergence of delta oscillations is also notable^9^ and might be related to the amplitude and spatial distribution of the ictal activity during the induced seizure (**Fig. 1**). However, oscillation changes in ECT and MST could not be directly compared here, due to the differences in data collection regimes between the two studies and the lack of data from cognitive and physical assessment. Further investigation is needed to more precisely describe the contributions of oscillations and aperiodic activity to the therapeutic and cognitive effects of ECT and MST.

Other important methodological limitations of this study include the fact that the MST cohort analyzed here has a 21% remission rate, which is notably lower than previous studies of MST^6,7^ that report remission rates as high as 50%. Thus, the MST cohort analyzed here might not reflect the general population. In addition, the majority of MST patients (61%) underwent 24 treatment sessions, which was the maximum number allowed according to the clinical protocol, thereby skewing the distribution of sessions needed to see a significant decrease in symptom severity. In contrast, the ECT dataset had a normal distribution of sessions before treatment cessation (See **Table 1, Supp. Fig. 3A,C**). This could be an effect of the two datasets being collected by different groups at different locations, thereby varying in clinical assessment, and physicians’ judgment on when to terminate treatment. Furthermore, the data in this interventional study and the preceding longitudinal study^29^ can only speak to the short-term effects of ECT and MST. More evidence is needed to determine how long-term changes in aperiodic and oscillatory activity persist post-treatment and if the longevity of these changes is linked to rates of MDD relapse. Lastly, ECT and MST post-treatment EEG recordings were collected at slightly different times. The ECT patients provided post-ECT EEG within 2 days after the last ECT session^35^, while MST patients waited longer on average to receive their post-MST EEG, with an average of 3.81 days after the last MST session^18^. Future research directly comparing ECT to MST must ensure to minimize methodological differences between treatment groups.

Future investigations can also explore the relationship of hemispheric differences and stimulation laterality to therapeutic efficacy and cognitive side effects. Because the vast majority of ECT and MST patients included in this study received bilateral stimulation and because we observed no regional specificity of spectral changes, our analyses included effects across both hemispheres. However, measures of hemispheric differences have been relevant to studies of depression, especially frontal alpha asymmetry, despite this measure being called into question by several recent meta-analyses^62,63^. More evidence is needed to uncover the role of hemispheric differences and regional specificity in general in aperiodic and periodic activity in ECT and MST.

The current study shows simultaneous, independent reductions in clinical severity and increases in aperiodic activity in MDD patients undergoing ECT or MST. Although not yet a viable clinical biomarker, the observed changes in aperiodic activity here provide further support for the cortical inhibition theory of depression^50^. Our results hint at a potential physiological interpretation of why ECT and MST are useful treatments for depression, though much more physiological evidence is required.

## Methods

### Summary of datasets

The data included in this analysis are from two previously conducted studies. The first, described in Voineskos et al., 2016^35^, is from an observational study of the EEG correlates of ECT efficacy for patients with MDD. This study was not a registered clinical trial. The second, described in Daskalakis et al., 2020^36^, is from a clinical trial studying MST for MDD, specifically as a cognition-sparing alternative to ECT (Clinicaltrials.gov, NCT01596608). The CONSORT flowchart for patients included in this study is available in supplemental materials. Both studies included resting-state EEG data and clinician-administered ratings of depression symptom severity collected first at baseline and again following treatment completion. The participants, treatment modalities, EEG data acquisition, and clinical measures for both the ECT study and MST clinical trial are described below.

### Participants

Twenty-two patients who received a diagnosis of MDD as per the Diagnostic and Statistical Manual (DSM-IV) were included in this study for ECT. Twenty-two patients with the same diagnosis were included in this study after participating in a clinical trial for MST (Clinicaltrials.gov, NCT01596608). Patients were considered as having treatment-resistant depression. See Table 1 for patient demographics and diagnoses. Ethnicity information was not collected at the time each study was conducted. Written informed consent was provided by all patients. Ethical approval was granted from the Centre for Addiction and Mental Health (CAMD) Research Ethics Board (REB) in accordance with the Declaration of Helsinki for both the ECT study (167-2009) and the MST clinical trial (145-2010). A complete list of inclusions and exclusions criteria is provided in Voineskos et al., 2016^35^ and Daskalakis et al., 2020^36^.

### Electroconvulsive therapy

Patients received ECT 2-3 times per week. Square wave pulses were delivered using an open label protocol with a brief-pulse device (MECTA Corporation, Lake Oswego, OR). Patients started their treatment with either right unilateral ultra-brief pulse width (0.3 msec) ECT, or brief pulse width (0.5-1.0 msec) bi- temporal ECT based on the preference of the treating physician and the patient. Electrode placement was in accordance with American Psychiatric Association guidelines. Patients who received unilateral treatment were later switched to bi-temporal ECT if they showed an initial poor response to treatment. Anesthesia was induced by administering methohexital for sedation, and succinylcholine for muscle relaxation. Treatment completion was based on clinical factors, patient response, the patient’s desire to discontinue treatment, or the most responsible physician’s clinical judgment. More details about this process can be found in Voineskos et al., 2016^35^.

### Magnetic seizure therapy

In a separate population participating in a clinical trial (Clinicaltrials.gov, NCT01596608), patients received MST 2-3 times per week. A twin coil (Twin Coil-XS) was used with a MagPro MST stimulator (Magvenure, Denmark). The two coils were placed bilaterally over the prefrontal cortex, approximating F3 and F4 locations (international 10-20 system). Anesthesia was induced by administering methohexital sodium or methohexital plus remifentanil for sedation, and succinylcholine for muscle relaxation. Treatment completion was based on clinical factors, defined as a remission if the HAMD-24 score < 10 and greater than 60% reduction in depressive symptoms using HAMD-24 scale, or a total of 24 treatments were administered. Notably, 61% of patients in this dataset received the maximum 24 treatments. More details can be found in Daskalakis et al., 2020^36^ and Hill et al., 2021^18^.

### Data acquisition

The data used in this study was previously acquired from Voineskos et al., 2016^35^, and Daskalakis et al., 2020^36^. EEG data were collected within a week before patients started their treatment. For the ECT dataset, post-ECT resting state EEG was collected within 2 days after their last treatment. Whereas for the MST dataset, post-MST resting state EEG were collected on average 3.81 days (SD 3.86)^18^ after their last treatment. A total of 10 minutes of resting state data with eyes closed were collected pre- and post-treatment for ECT and MST datasets. A 64-electrode cap (Neuroscan *Quik-Cap*) containing sintered Ag/AgCl electrodes connected to a SynAmps^2^ amplifier (Neuroscan, Compumedics, USA) was used for all recordings (online reference and ground electrodes located at the vertex, and just posterior to Fz, respectively). Impedances were required to remain below 5kΩ prior to starting the recording, and data quality was monitored throughout the session. The sampling frequency was either 1000Hz or 10,000Hz. More details can be found in Voineskos et al., 2016^35^ and Daskalakis et al., 2020^36^.

### Clinical measures

Demographic and medication information were recorded at baseline during clinical interview (**Table 1**). For the ECT dataset, the primary clinical measure was the 17-item Hamilton Depression Rating Scale (HAMD-17), which was completed before the first treatment sessions, and within 2 days after the last. Of the 22 patients who had complete pre- and post-ECT EEG recordings, 19 had complete pre- and post-ECT HAMD-17 ratings. For the MST dataset, the 24-item HAM-D was used and of the 22 patients who had complete pre- and post-MST EEG recordings, 14 had complete pre- and post-MST HAMD-24 ratings. Because these datasets were collected independently, and using different versions of the HAM-D, clinical improvement was assessed separately for ECT and MST. We calculated remission rate as the percentage of the included patients that showed equal or larger than 50% decrease in symptom severity.

### EEG pre-processing

Patients whose EEG data was recorded with a sampling frequency of 10,000Hz were downsampled to 1000Hz. Then bad electrodes were identified and removed based on the presence of excess noise by inspecting the raw time series and the power spectra per electrode. After this, the data were re-referenced to the common average. In order to improve the performance of ICA, a FIR high-pass filter of 0.5 Hz was applied with a Hamming window. Fast ICA was used with 15 components to remove eye movements, eye blinks, and other non-neural artifacts. Twenty components were used for data that had considerable artifacts, which was the case for 4 ECT recordings and 5 MST recordings. ICA was then applied to the raw, unfiltered data to avoid distorting spectral power <0.5 Hz. Lastly, the bad electrodes were interpolated using the spherical spline interpolation method implemented with the MNE function raw.interpolate_bads().

### EEG Spectral Parameterization

Power spectra were computed per patient for each electrode from the continuous EEG data using Welch’s method, with a Hamming window of 4 seconds, and 2 second overlap between windows.

The spectral parameterization model was fit to each power spectrum between 0.5 and 30 Hz, without a knee, and oscillation peaks were defined as “bumps” that surpassed a threshold of 0.05 µV^2^ above the aperiodic component. A maximum of 12 peaks were fit with a minimum band width of 1 Hz and maximum of 8 Hz. On average, three peaks were found per power spectrum, both pre- and post-treatment. Furthermore, the three frequency ranges of interest were delta (1-4 Hz), theta (4-7 Hz), and alpha (7-12 Hz). The peak with the highest power was selected within each frequency range. Model fits (R^2^) below 0.8 were excluded from further analysis; data from that electrode in corresponding post- (or pre-) recording was also removed. One patient in the ECT dataset and one patient in the MST dataset were completely removed from further analysis, due to excessive noise over the majority of the electrodes, which caused systematic model fits below the threshold. Overall, model fits were excellent with an average of 1.4 electrodes dropped per ECT patient, and 0.4 electrodes dropped per MST patient. Canonical band power was calculated in addition to the metrics extracted using spectral parameterization methods for the purpose of comparing methodological approaches for quantifying parameterized spectral power to traditional band power. Band power was computed as the mean of spectral power in each frequency band (delta, theta, and alpha).

Although our previous exploratory results were restricted to frontal electrodes^29^ and much prior research has identified frontal regions as the site of the strongest effects of MDD treatment^64,65^, we decided to include all scalp electrodes in this analysis. Scalp topographies did not suggest a characteristic distribution of EEG changes. In order to thoroughly investigate the electrophysiological drivers of clinical slowing, no subset of electrodes were favored or excluded from analysis based on topography. Furthermore, we calculated the fraction of all electrodes containing an oscillation peak per frequency band of interest (termed oscillation abundance) in order to capture the emergence of oscillations, which might contribute to observations of clinical slowing. Importantly this emergence is different from merely an increase in the power of existing oscillations. Overall, features of interest considered for analysis from the EEG signal included the aperiodic exponent, delta oscillation power, delta band power, delta abundance, theta oscillation power, theta band power, theta abundance, alpha oscillation power, alpha band power, and alpha abundance. Prior to statistical analysis and visualization, features of interest were averaged across all included EEG electrodes for each patient. All difference values were calculated as pre-minus post-.

### Statistical Analyses

#### Multiple regression – relating band power to aperiodic exponent and oscillations

Band power is traditionally used to compute power within certain frequency ranges. Therefore, we wanted to include an ordinary least squares regression to see whether it is the aperiodic exponent, actual oscillation power within a frequency range, or abundance of oscillations across the scalp that is more associated with these band power measures. We did this for three frequency ranges of interest: delta, theta and alpha. For these regressions we used the pre- minus post- differences in exponent, oscillation power, and oscillation abundance. Thus, the formula for delta band power is

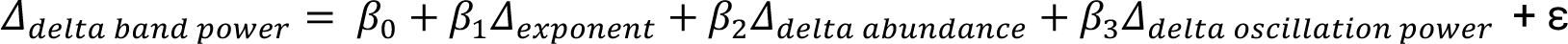

The formula for theta band power is

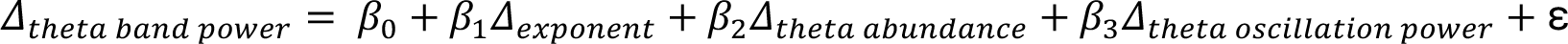

Last, the formula for alpha band power is

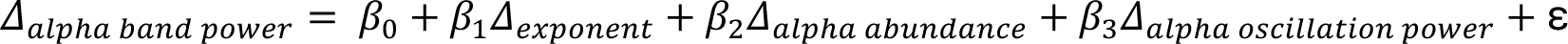

To adjust our regressions for Type I inflation, we reduced the ɑ-threshold according to a Holm-Bonferroni correction. Therefore, the lowest ɑ-threshold for the three main effects in the model is 0.0167 (e.g., 1.67 x 10^-2^) instead of the conventional 0.05. Standardized main effects coefficients for each regression were reported with corresponding p-values and 95% confidence intervals.

#### Multiple regression – Relating clinical outcome to EEG spectral parameters

To assess whether any of the observed changes or baseline EEG features could be associated with clinical outcome across both ECT and MST treatment groups, we performed an ordinary least squares regression. In order to identify potential physiological indicators viable for both ECT and MST treatment, we combined the two datasets. In order to combine the datasets, we normalized the HAM-D scores from the HAMD-17 used in the ECT study and the HAMD-24 used in the MST trial onto a single, linearized 0-1 scale using the following formula. Here, the maximum score for the HAMD-17 was 52 and the maximum score for HAMD- 24 was 76.

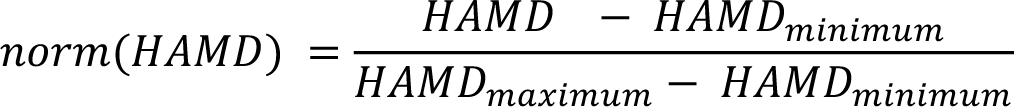

To select features to include in the model, we performed an exhaustive search (Exhaustive Feature Selector from mlxtend), where normalized post-treatment HAM-D score was predicted with two fixed effects, pre-treatment HAM-D score and a categorical variable of treatment type, and at least one other main effect, which was determined by the exhaustive search algorithm. This other main effect was selected from the following features: baseline exponent, baseline delta abundance, baseline theta abundance, change in exponent, change in delta abundance, change in theta abundance, and number of treatments received. These features were chosen for exhaustive search in order to consider traditional measures of clinical slowing in delta and theta frequency bands in addition to new measures of aperiodic activity. Number of treatments was included to account for the dose-response effect of multiple treatments. Using R^2^adj for model evaluation, the exhaustive search algorithm identified the following formula to best predict normalized post-treatment HAM-D score across both ECT and MST datasets. Of the potential features included in the search, baseline exponent demonstrated the strongest relationship to clinical outcome:

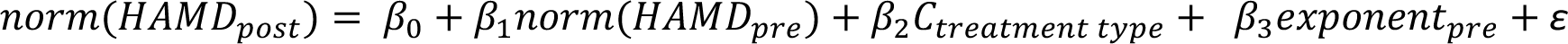

Furthermore, in running the multiple regression following the above formula, we used an HC3 heteroskedasticity-consistent covariance matrix to account for heteroskedasticity in the data. The HC3 correction has been shown to work best for sample sizes <250^66^. Standardized main effect coefficients were reported with corresponding p-values and 95% confidence intervals.

#### Observational difference tests

To test statistical significance between pre- and post-treatment clinical outcomes, we have used a Wilcoxon signed rank test instead of a paired t-test, since the data was not normally distributed. Cliff’s Delta was used to measure effect size, where 0.147 is a small effect, 0.33 is a medium effect, and 0.47 is large^67^.

The dependent variables considered for analysis from the EEG signal are the aperiodic exponent, delta oscillation power, delta band power, delta abundance, theta oscillation power, theta band power, theta abundance, alpha oscillation power, alpha band power, and alpha abundance. All of these parameters were calculated before the treatment started (pre), and after the treatment was completed (post). Normality was checked using the Shapiro-Wilk test. A two-sided paired t-test was performed on the pre- vs post-variables if the data were normally distributed. Otherwise, a Willcoxon signed rank test was used. If paired t-tests were performed, the mean and standard deviations were reported, and Cohen’s d was used to determine effect size. If the Wilcoxon signed rank test was performed, the median and interquartile range were reported, and Cliff’s Delta was used to determine effect size.

Importantly, to correct for multiple comparisons and Type I error, we implemented Holm-Bonferroni corrections. Within ECT and MST datasets, we conduct eight tests on the EEG features. Therefore, our ɑ-threshold is adjusted based on the number of hypothesis tests performed and the rank of each test’s p-value, instead of the conventional 0.05. Adjusted alpha values are included with results statistics and are available in **Supp. Table 1**.

### Software

All EEG (pre-)processing, statistical analyses, and plotting was performed in Python (3.9.7), using MNE 0.24.1)^68^, Spectral Parameterization (FOOOF; 1.0.0)^24^, Pandas (1.3.2)^69^, Pingouin (0.5.0)^70^, Statsmodels (OSL) (0.13.1)^71^, mlxtend (0.22.0)^72^, Matplotlib (3.4.2)^73^, numpy (1.22.4)^74^, scipy (1.7.1)^75^, and Seaborn (0.11.2)^76^.

## Data availability

All code used for all analyses and plots are publicly available on GitHub at https://github.com/voytekresearch/ect-mst. The data collected in this study is not available at this time.

## Data Availability

Data presented in the study is not available at this time.

## Acknowledgements

Support: NIH National Institute of General Medical Sciences grant R01GM134363-01 (to B.V.)

Thanks: We thank Andrew Bender, Dillan Cellier, Ryan Hammonds, Blanca Martin-Burgos, Michael Preston, and Trevor McPherson for their advice and feedback on the manuscript.

## Author contributions

S.E.S., I.H., and B.V. conceived of the experiments and developed the analyses. I.H., R.Z., A.T.H., Z.J.D., and D.M.B., collected the data. S.E.S., E.L.K., Q.vE., and B.V. wrote analysis code and analyzed the data. J.K. provided essential support for statistical analysis. S.E.S., E.L.K., Q.vE., I.H., and B.V. wrote the manuscript, and all authors edited the manuscript.

**Competing interests** A.T.H. was supported by an Alfred Deakin Postdoctoral Research Fellowship. D.M.B. receives research support from the Canadian Institutes of Health Research (CIHR), National Institutes of Health – US (NIH), Brain Canada Foundation and the Temerty Family through the CAMH Foundation and the Campbell Family Research Institute. He received research support and in-kind equipment support for an investigator-initiated study from Brainsway Ltd. and he was the site principal investigator for three sponsor-initiated studies for Brainsway Ltd. He received in-kind equipment support from Magventure for investigator-initiated studies. He received medication supplies for an investigator-initiated trial from Indivior. He has participated in an advisory board for Janssen. He has participated in an advisory board for Welcony Inc. Z.J.D. has received research and equipment in-kind support for an investigator-initiated study through Brainsway Inc and Magventure Inc and industry-initiated trials through Magnus Inc. He also currently serves on the scientific advisory board for Brainsway Inc. His work has been supported by the National Institutes of Mental Health (NIMH), the Canadian Institutes of Health Research (CIHR), Brain Canada and the Temerty Family, Grant and Kreutzcamp Family Foundations.

## Notes

### Competing Interest Statement

ATH was supported by an Alfred Deakin Postdoctoral Research Fellowship. DMB receives research support from the Canadian Institutes of Health Research (CIHR), National Institutes of Health - US (NIH), Brain Canada Foundation and the Temerty Family through the CAMH Foundation and the Campbell Family Research Institute. He received research support and in-kind equipment support for an investigator-initiated study from Brainsway Ltd. and he was the site principal investigator for three sponsor-initiated studies for Brainsway Ltd. He received in-kind equipment support from Magventure for investigator-initiated studies. He received medication supplies for an investigator-initiated trial from Indivior. He has participated in an advisory board for Janssen. He has participated in an advisory board for Welcony Inc. ZJD has received research and equipment in-kind support for an investigator-initiated study through Brainsway Inc and Magventure Inc and industry-initiated trials through Magnus Inc. He also currently serves on the scientific advisory board for Brainsway Inc. His work has been supported by the National Institutes of Mental Health (NIMH), the Canadian Institutes of Health Research (CIHR), Brain Canada and the Temerty Family, Grant and Kreutzcamp Family Foundations.

### Author Declarations

Ethics Committee of the Center for Addiction and Mental Health gave ethical approval for the study in accordance with the declaration of Helsinki.

### Summary of Updates

Title and abstract updated to incorporate entire scalp analysis instead of frontal only Results and statistics updated to correct for multiple comparisons, add exhaustive search and multiple linear regression. Language broadly updated to reflect new results.

