## Supplementary Material for "Magnetic seizure therapy and electroconvulsive therapy increase aperiodic activity"

ECT: In the 20 ECT patients who exhibited theta oscillations both pre- and post-ECT, the regression to predict theta band power was overall significant ($R^2_{adj}$ = 0.74, F(3, 16) = 18.90, p = 1.64 x $10^{-5}$). An increase in aperiodic exponent does not predict theta band power (β = 0.25, $\alpha_{adj}$ = 2.50 x $10^{-2}$, p = 0.06, 95% CI[-0.01, 0.52]), nor does theta abundance (β = -0.01, $\alpha_{adj}$ = 0.05, p = 0.96, 95% CI[-0.27, 0.25]), but theta oscillation power was significantly related to theta band power (β= 0.77, $\alpha_{adj}$ = 1.67 x $10^{-2}$, p = 1.80 x $10^{-5}$, 95% CI[0.50, 1.05]). All 22 patients exhibited alpha oscillations both pre- and post-ECT, and the overall regression to predict alpha band power was significant ($R^2_{adj}$ = 0.50, F(3, 18) = 7.89, p = 1.44 x $10^{-3}$). Alpha band power is significantly related to alpha aperiodic adjusted power (β = 0.65, $\alpha_{adj}$ = 1.67 x $10^{-2}$, p = 1.2 x $10^{-3}$, 95% CI[0.29, 1.00]), but not by alpha abundance (β = -0.31, $\alpha_{adj}$ = 2.50 x $10^{-2}$, p = 0.08, 95% CI[-0.66,0.04]), nor a change in exponent (β = 0.16, $\alpha_{adj}$ = 0.05, p = 0.37, 95% CI[-0.20, 0.52]).

MST: In the 18 patients who exhibited theta oscillations both pre- and post-MST, the regression to predict theta band power was overall significant ($R^2_{adj}$ = 0.89, F(3, 14) = 47.33, p = 1.42 x $10^{-7}$). Theta band power was significantly related to both a change in exponent (β = 0.42, $\alpha_{adj}$ = 2.50 x $10^{-2}$, p = 1.73 x $10^{-3}$, 95% CI[0.19, 0.67]) and theta oscillation power (β = 0.65, $\alpha_{adj}$ = 1.67 x $10^{-2}$, p = 1.01 x $10^{-4}$, 95% CI[0.39, 0.91]). Theta abundance was not related to theta band power (β = -0.11, $\alpha_{adj}$ = 0.05, p = 0.25, 95% CI[-0.30, 0.09]). All 22 patients exhibited alpha oscillations both pre- and post-MST, and the regression for alpha band power was overall significant ($R^2_{adj}$ = 0.45, F(3, 18) = 6.63, p = 3.29 x $10^{-3}$). Both aperiodic exponent (β = 0.64, $\alpha_{adj}$ = 2.50 x $10^{-2}$, p = 4.57 x $10^{-3}$, 95% CI[0.23, 1.06]) and aperiodic adjusted alpha power (β = 0.85, $\alpha_{adj}$ = 1.67 x $10^{-2}$, p = 4.22 x $10^{-4}$, 95% CI[0.44, 1.27]) were significantly related to alpha band power, but not alpha abundance (β = 0.06, $\alpha_{adj}$ = 0.05, p = 0.73, 95% CI[-0.29, 0.41]).

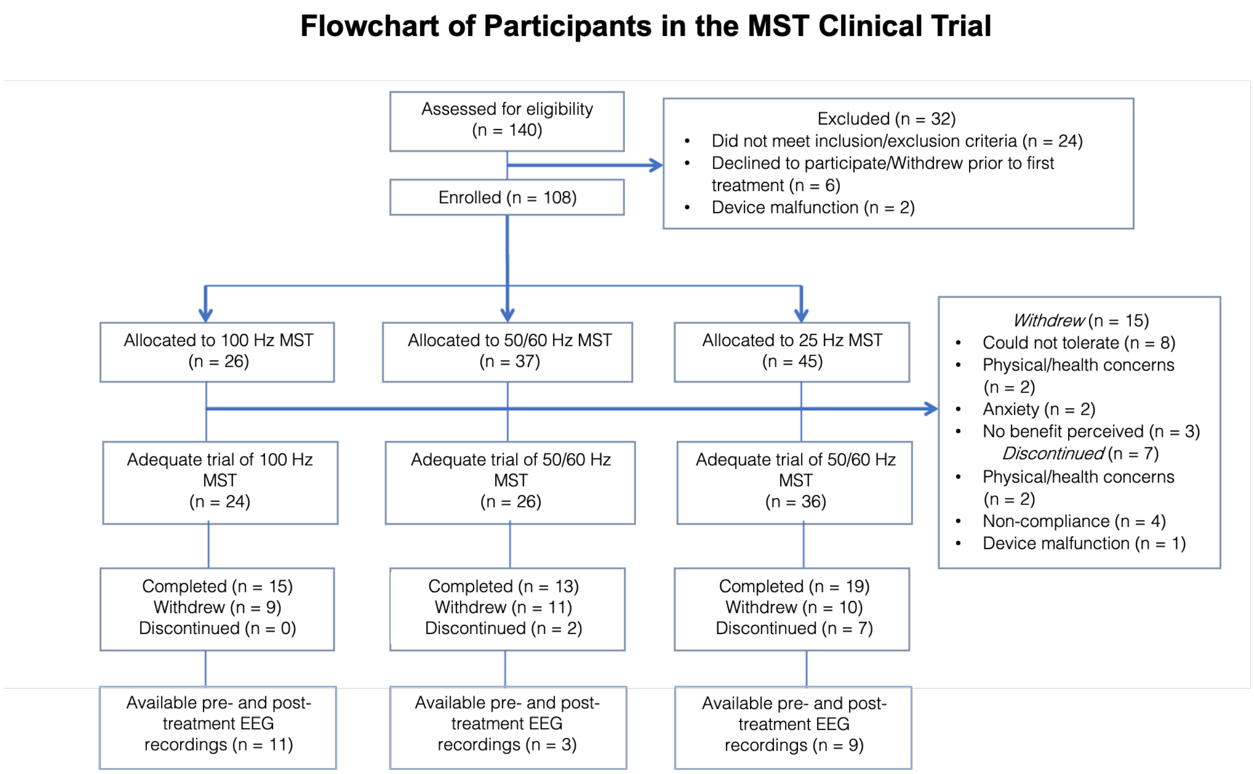


**Supplementary Fig. 1: CONSORT flowchart**

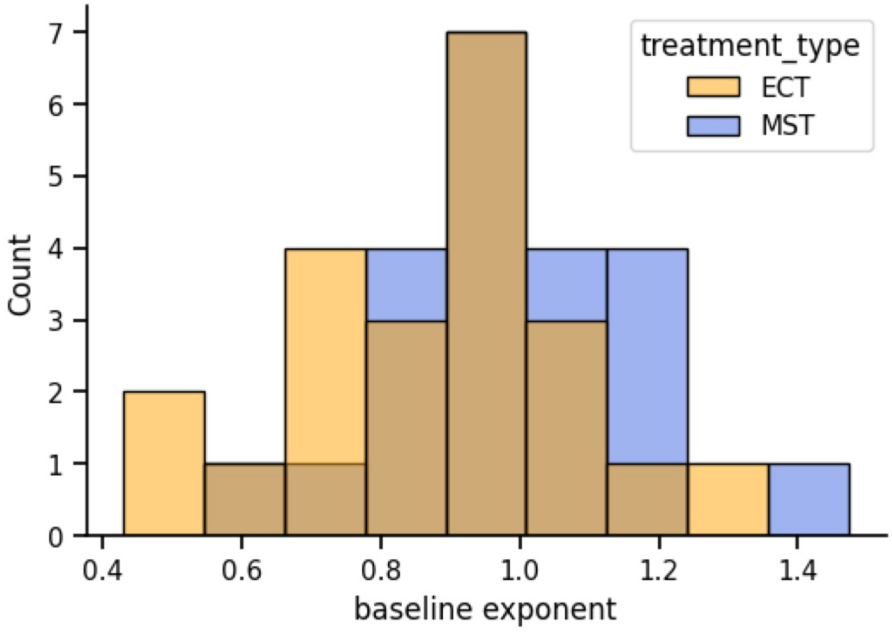


**Supplementary Fig. 2: Baseline differences in EEG aperiodic exponent of patients receiving ECT and MST.** There is no significant difference in baseline aperiodic exponent between patients receiving either ECT or MST (ECT = 0.88 ± 0.21, MST = 0.98 ± 0.18, $t(42) = 1.71$, $p = 0.094$).

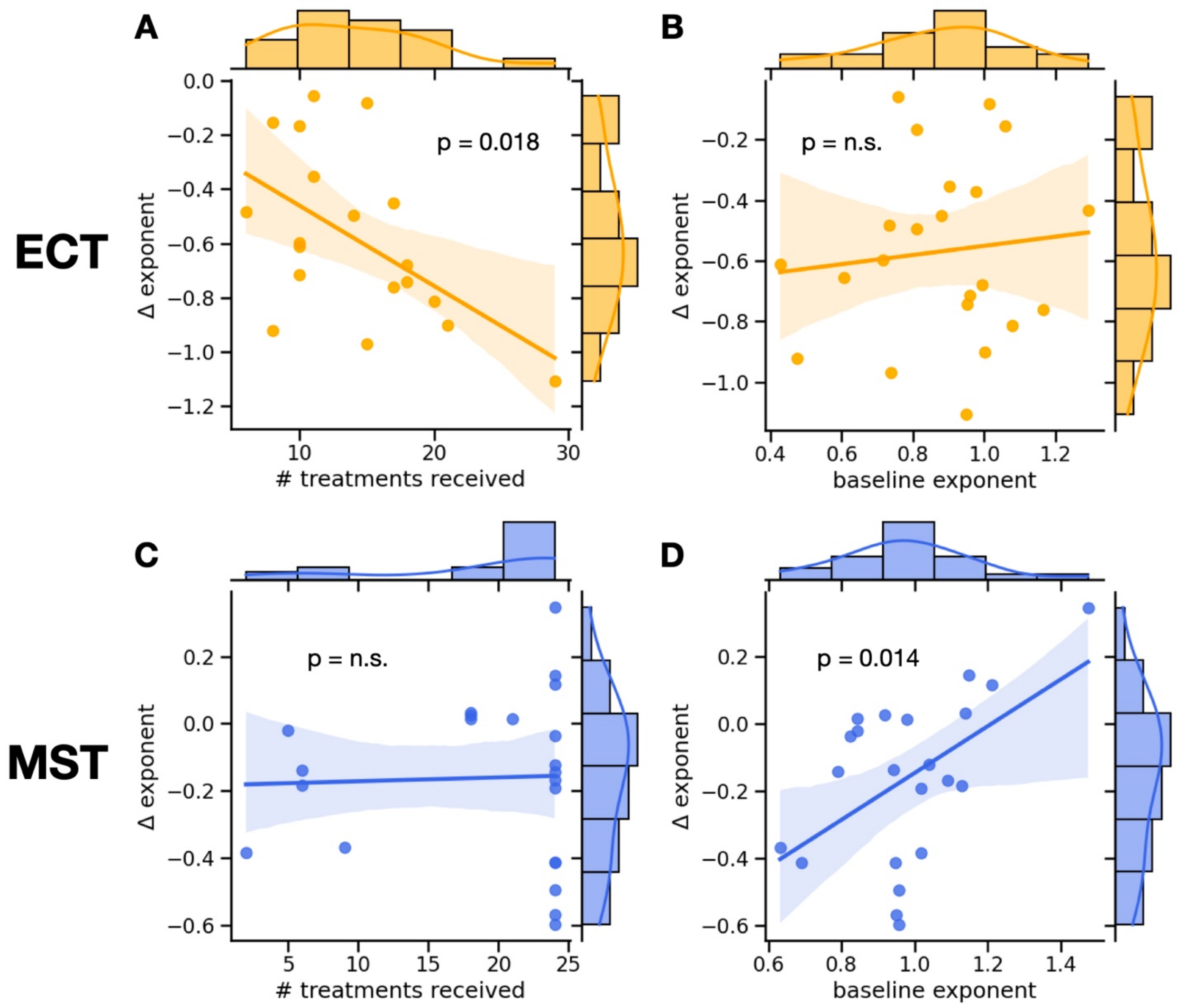


**Supplementary Fig.3: Correlations between the difference in exponent due to ECT or MST and the number of treatments received or the baseline exponent.** In ECT, the number of treatments received is significantly negatively correlated to the difference in exponent (r = -0.54, p = 0.018). Thus, the more ECT treatments received, the "steeper" the spectrum becomes. Whereas the baseline exponent is not correlated to the difference in exponent (r = 0.11, p = 0.63). In MST, the number of treatments received is not correlated to the difference in exponent (r = 0.034, p = 0.87). Whereas the baseline exponent is significantly positively correlated to the difference in exponent (r = 0.51, p = 0.014). Thus, the "flatter" the spectrum at baseline, the more "steepening" we see due to MST.

**Supplementary Table 1: Holm-Bonferroni correction values.** Overview of multiplicity-correction for ECT and MST datasets. Each dataset had 8 hypothesis tests performed on EEG features. Holm's Sequential Bonferroni procedure was applied to adjust alpha threshold for significance testing.

| treatment type | Feature for hypothesis test | p-value | rank | alpha (adjusted) | significant? |
|---|---|---|---|---|---|
| ECT | exponent | 1.05E-08 | 1 | 6.25E-03 | yes |

| | | | | | |
|---|---|---|---|---|---|
| | delta band power | 4.83E-08 | 2 | 7.14E-03 | yes |
| | delta oscillation power | 2.44E-02 | 8 | 5.00E-02 | yes |
| | delta abundance | 1.77E-04 | 4 | 1.00E-02 | yes |
| | theta oscillation power | 1.90E-05 | 3 | 8.33E-03 | yes |
| | theta abundance | 5.40E-03 | 6 | 1.67E-02 | yes |
| | alpha oscillation power | 3.20E-03 | 5 | 1.25E-02 | yes |
| | alpha abundance | 1.90E-02 | 7 | 2.50E-02 | yes |
| MST | exponent | 6.00E-03 | 2 | 7.14E-03 | yes |
| | delta band power | 3.65E-02 | 5 | 1.25E-02 | no |
| | delta oscillation power | 9.80E-03 | 3 | 8.33E-03 | no |
| | delta abundance | 7.96E-01 | 8 | 5.00E-02 | no |
| | theta oscillation power | 3.80E-05 | 1 | 6.25E-03 | yes |
| | theta abundance | 1.81E-02 | 4 | 1.00E-02 | no |
| | alpha oscillation power | 3.91E-01 | 7 | 2.50E-02 | no |
| | alpha abundance | 1.78E-01 | 6 | 1.67E-02 | no |
